# Impact of low relative skeletal muscle mass and power on the development of metabolic syndrome in Japanese women: a 7-year prospective study

**DOI:** 10.1101/2022.01.27.22269435

**Authors:** Yosuke Yamada, Haruka Murakami, Ryoko Kawakami, Yuko Gando, Hinako Nanri, Takashi Nakagata, Daiki Watanabe, Tsukasa Yoshida, Yoichi Hatamoto, Eiichi Yoshimura, Kiyoshi Sanada, Nobuyuki Miyatake, Motohiko Miyachi

**Author notes:** Correspondence: Yosuke Yamada, PhD, National Institute of Health and Nutrition, National Institutes of Biomedical Innovation, Health and Nutrition, 1-23-1, Toyama, Shinjuku-ku, Tokyo 162-8636, Japan.

## Abstract

Previous cross-sectional studies indicated that low relative skeletal muscle mass against body weight (appendicular lean mass divided by body weight, ALM/Wt or divided by body mass index, ALM/BMI) was negatively associated with metabolic syndrome (MetS). The aim of this longitudinal study was to investigate the impact of low relative skeletal muscle mass and leg muscle power on the development of MetS in Japanese women in a 7-y prospective study. Subjects were 346 Japanese women aged 26 to 85 years. The reference values for Class I sarcopenia were defined as values one SD below the sex-specific means of the young group. The longitudinal relation between muscle mass or leg power and development of MetS was examined using Kaplan-Meier curves and Cox regression models (average follow-up duration 7 years, range 1 to 10 years). During follow-up, 24 of subjects developed MetS. Both indices of ALM/Wt or ALM/BMI, the incidence of MetS was higher in Class I sarcopenia group than normal muscle mass group even after controlling variables (adjusted hazard ratio [AHR] = 3.22 [95%CI; 1.25-8.27] for ALM/Wt and 3.19 [95%CI; 1.31-7.74] for ALM/BMI). In contrast, for the leg power per body weight, the incidence of MetS was not significantly different between low leg power and normal leg power groups (AHR = 1.23 [95%CI; 0.50-3.04]). Current longitudinal study indicated that low relative skeletal muscle mass but not low leg power was independent risk factors for the development of MetS in Japanese women.

## 1. Introduction

Skeletal muscle mass (SMM) and its function are considered important biomarkers of aging [1-3]. Age-related loss of SMM and its function is called as sarcopenia [4]. In current measureable definition of sarcopenia is based on SMM and grip strength and/or lower body physical performance [1, 2]. Skeletal muscle is a metabolically active organ to mediate energy metabolism and exert beneficial effects on metabolic health [5, 6]. Thus, higher muscle mass or muscle function might have beneficial effect on preventing metabolic syndrome (MetS) as well [7-11]. MetS is a cluster of conditions that occur together, increasing the risk of heart disease, stroke and type 2 diabetes. These conditions include the accumulation of visceral fat, increased blood pressure, dyslipidemia, and hyperglycemia.

However, because the people who have higher body weight tend have higher muscle mass and strength, previous studies indicated that the normalization of muscle mass and strength by body weight or body mass index (BMI) is needed to see the association between skeletal muscle mass or strength and MetS [7-14]. Previous cross-sectional studies indicated that relative appendicular lean mass per kg weight or per body mass index is inversely associated with MetS [7-14]. The present longitudinal study aimed to examine the impact of low relative skeletal muscle mass and power on the development of MetS in Japanese women.

## 2. Materials and Methods

### 2.1. Ethics approval and consent to participate

The study was performed in accordance with the guidelines of the Declaration of Helsinki. All procedures were reviewed and approved by the ethics committees of the National Institutes of Biomedical Innovation, Health and Nutrition (6008, Kenei14-02). All participants provided written consent for participation in the study.

### 2.2. Participants

The participants were recruited from the community around the National Institute of Health and Nutrition, Tokyo, Japan. From a total of 760 women, 346 women aged 26 to 85 years old (mean and SD of age, ± years) were included in the current study who met the following criteria: (1) they received anthropometric and physical activity measurements, (2) underwent blood examinations, (3) dietary intake assessments, (4) no history of MetS at the baseline measurement, and (5) underwent follow-up examinations.

### 2.3. Anthropometric and leg power measures

Height, weight, and waist circumference were measured and body mass index was calculated. Body composition was determined by bioelectrical impedance analysis (TANITA BC-600). The appendicular lean mass (ALM; kg) were obtained from the sum of the four limbs (right and left upper and lower limbs). The skeletal muscle index (SMI) was calculated as ALM divided by body weight × 100 (%). The reference values for Class I sarcopenia were defined as values one SD below the sex-specific means of the young group, and 28.0% was the cutoff value for Class I sarcopenia. In addition, ALM divided by BMI (ALM/BMI) was also calculated based on FNIH recommendation. The cutoff value for Class I sarcopenia of ALM/BMI was 0.670. We used these values to divide the subject into normal muscle mass group and Class I sarcopenia group. No women fell into Class II sarcopenia in the current subjects.

The leg extension power was measured by using a dynamometer (Anaero Press 3500; Combi Wellness, Tokyo, Japan) in the sitting position [15]. The subjects were advised to vigorously extend their legs. 5 trials were performed at 15-s intervals, and the average of the 2 highest recorded power outputs (watt; W) was taken as the definitive measurement. The leg extension power divided by body weight was obtained.

### 2.4. Physical Activity

The duration and intensity of physical activity were evaluated by a triaxial accelerometer (Actimarker EW4800; Panasonic, Osaka, Japan) [16, 17], as described previously [15]. Participants were asked to wear the physical activity monitor on their hip for 28 days; we used data for 14 days, during which the accelerometer was worn continuously from the time the participant awoke until she went to bed. Total volume of physical activity (METs × hour) was obtained.

### 2.5. Blood samples

Blood samples were taken from participants following an overnight fast of at least 10 hours [18]. Venous blood withdrawn from the antecubital vein was collected into tubes without additives or EDTA and was immediately centrifuged at 3000 rpm for 20 minutes to obtain serum or plasma. The levels of glucose, HbA1c, homeostasis model assessment of insulin resistance (HOMA-IR), and HOMA-β in plasma and total cholesterol, high-density and low-density lipoprotein (HDL and LDL) cholesterol, and triglycerides in serum were determined [15].

According to the definition released by the Japanese Committee for the Diagnostic Criteria of Metabolic Syndrome in April 2005, we defined metabolic syndrome as the presence of 2 or more abnormalities in addition to visceral obesity (waist circumference: 85 cm or more in men, 90 cm or more in women). These three abnormalities are as follows: 1, triglycerides ≥150 mg/dL and/or HDL-cholesterol <40 mg/dL or under treatment for this type of dyslipidemia, 2, systolic blood pressure ≥130 and/or diastolic blood pressure ≥85, or under treatment for hypertension, 3, fasting glucose ≥110 mg/dL or under treatment for diabetes [19].

### 2.6. Statistical analysis

Results are presented as mean ± standard deviation (SD). Differences were analyzed using ANOVA. Cumulative event rates for incident MetS were estimated by Kaplan-Meier curves, and the equalities were compared with the log-rank test. Cox proportional hazard analysis was performed to determine the independent association between either baseline SMI, ALM/BMI, or leg power against other variables. For multivariate analysis, model 1 was a crude form; age was adjusted for in model 2; model 3 included model-2 adjustment and family history of diabetes, smoking status, and physical activity. Alpha of 0.05 was employed to denote significant statistical deviation. We performed all analyses using IBM SPSS Statistics for Windows, version 22.0 (IBM Corp., Armonk, NY).

## 3. Results

Table 1 shows the baseline characteristics of the subjects according to SMI group. The subjects with Class I sarcopenia had higher weight, waist circumference, fasting glucose, HOMA-IR, SBP, DBP, triglycerides, and LDL cholesterol compared with normal SMI subjects. In contrast, the subjects with Class I sarcopenia had lower SMI, ALM/BMI, and leg power compared with normal SMI subjects.

**Table 1.** Baseline characteristics of the study subjects according to SMI group (N = 346)

| SMI (%) | Normal SMI |  |  |  |  |  | Class I sarcopenia |  |  | P value |
| --- | --- | --- | --- | --- | --- | --- | --- | --- | --- | --- |
|  | Above Ave. |  |  | Below Ave. |  |  |  |  |  |  |
|  | ≥32% |  |  | 28%-32% |  |  | <28% |  |  |  |
|  | n = 81 |  |  | n = 184 |  |  | n = 81 |  |  |  |
|  | mean | ± | SD | mean | ± | SD | mean | ± | SD |  |
| Age (y) | 54.1 | ± | 11.7 | 56.6 | ± | 11.6 | 55.7 | ± | 9.3 | 0.24 |
| Weight (kg) | 48.5 | ± | 5.0 | 53.9 | ± | 5.3 | 62.0 | ± | 7.4 | <0.001 |
| BMI (kg/m<sup>2</sup>) | 19.7 | ± | 1.5 | 21.9 | ± | 2.1 | 25.2 | ± | 2.8 | <0.001 |
| Waist circumference (cm) | 73.3 | ± | 6.2 | 80.4 | ± | 7.1 | 90.3 | ± | 7.1 | <0.001 |
| SMI (%) | 33.4 | ± | 1.4 | 30.0 | ± | 1.1 | 26.5 | ± | 1.1 | <0.001 |
| ALM/BMI | 0.825 | ± | 0.079 | 0.740 | ± | 0.060 | 0.655 | ± | 0.062 | <0.001 |
| HbA1c (%) | 5.4 | ± | 0.5 | 5.5 | ± | 0.4 | 5.6 | ± | 0.5 | 0.115 |
| Fasting glucose (mg/dL) | 89.2 | ± | 14.2 | 89.6 | ± | 11.1 | 94.0 | ± | 13.0 | 0.015 |
| HOMA-IR | 0.69 | ± | 0.38 | 0.86 | ± | 0.36 | 1.39 | ± | 1.16 | <0.001 |
| SBP (mmHg) | 114.2 | ± | 18.3 | 120.0 | ± | 16.8 | 120.0 | ± | 16.8 | 0.008 |
| DBP (mmHg) | 67.9 | ± | 10.8 | 71.3 | ± | 10.4 | 72.1 | ± | 10.1 | 0.020 |
| HR (bpm) | 58.2 | ± | 8.3 | 59.8 | ± | 9.3 | 61.3 | ± | 8.7 | 0.093 |
| Total cholesterol (mg/dL) | 210.0 | ± | 37.7 | 221.1 | ± | 35.4 | 219.4 | ± | 30.9 | 0.055 |
| Triglycerides (mg/dL) | 70.1 | ± | 31.4 | 83.0 | ± | 43.1 | 104.3 | ± | 55.8 | <0.001 |
| HDL cholesterol (mg/dL) | 75.5 | ± | 14.7 | 72.7 | ± | 17.8 | 62.9 | ± | 11.8 | <0.001 |
| LDL cholesterol (mg/dL) | 120.1 | ± | 32.0 | 131.4 | ± | 30.4 | 135.3 | ± | 28.3 | 0.004 |
| PA (METs*hour) | 4.4 | ± | 2.9 | 3.8 | ± | 1.7 | 3.8 | ± | 1.9 | 0.077 |
| Leg Power (W/kg) | 16.3 | ± | 3.4 | 14.7 | ± | 4.1 | 13.8 | ± | 3.8 | <0.001 |

SMI, skeletal muscle mass index; BMI, body mass index; ALM, appendicular lean mass; SBP, systolic blood pressure; DBP, diastolic blood pressure; HR, heart rate; HDL, high density lipoprotein; LDL, low density lipoprotein; PA, physical activity.

Kaplan-Meier curves for events of incident MetS according to baseline SMI. Fig 1 shows the Kaplan-Meier curves for events of incident MetS according to baseline SMI, and the subjects with Class I sarcopenia defined by SMI had significantly higher incident of MetS during follow-up period (P = 0.018). Fig. 2 shows the subjects with Class I sarcopenia defined by ALM/BMI had also significantly higher incident of MetS (P < 0.001). In contrast, high ALM/height^2^ or high absolute ALM had significantly higher incident of MetS (P < 0.001). Fig. 3 shows the Kaplan-Meier curves for events of incident MetS according to baseline leg power, and the subjects with low leg power had significantly higher incident of MetS (P = 0.032).

**Fig. 1.**
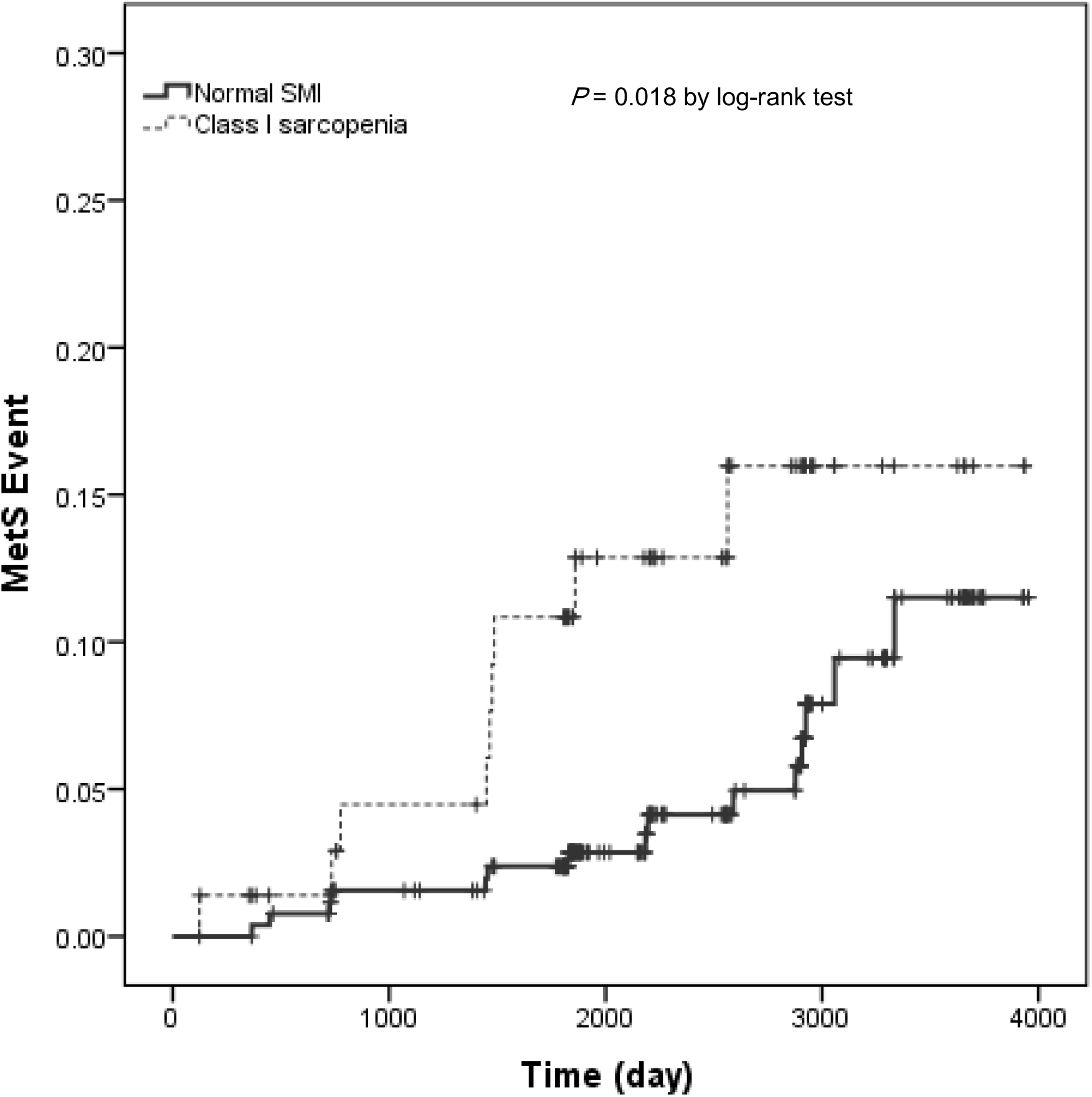
Kaplan-Meier curves for events of incident metabolic syndrome (MetS) according to baseline skeletal muscle index (SMI, %). Bold line shows normal SMI, and dashed line shows Class I sarcopenia. The SMI was calculated as ALM divided by body weight × 100 (%).

**Fig. 2.**
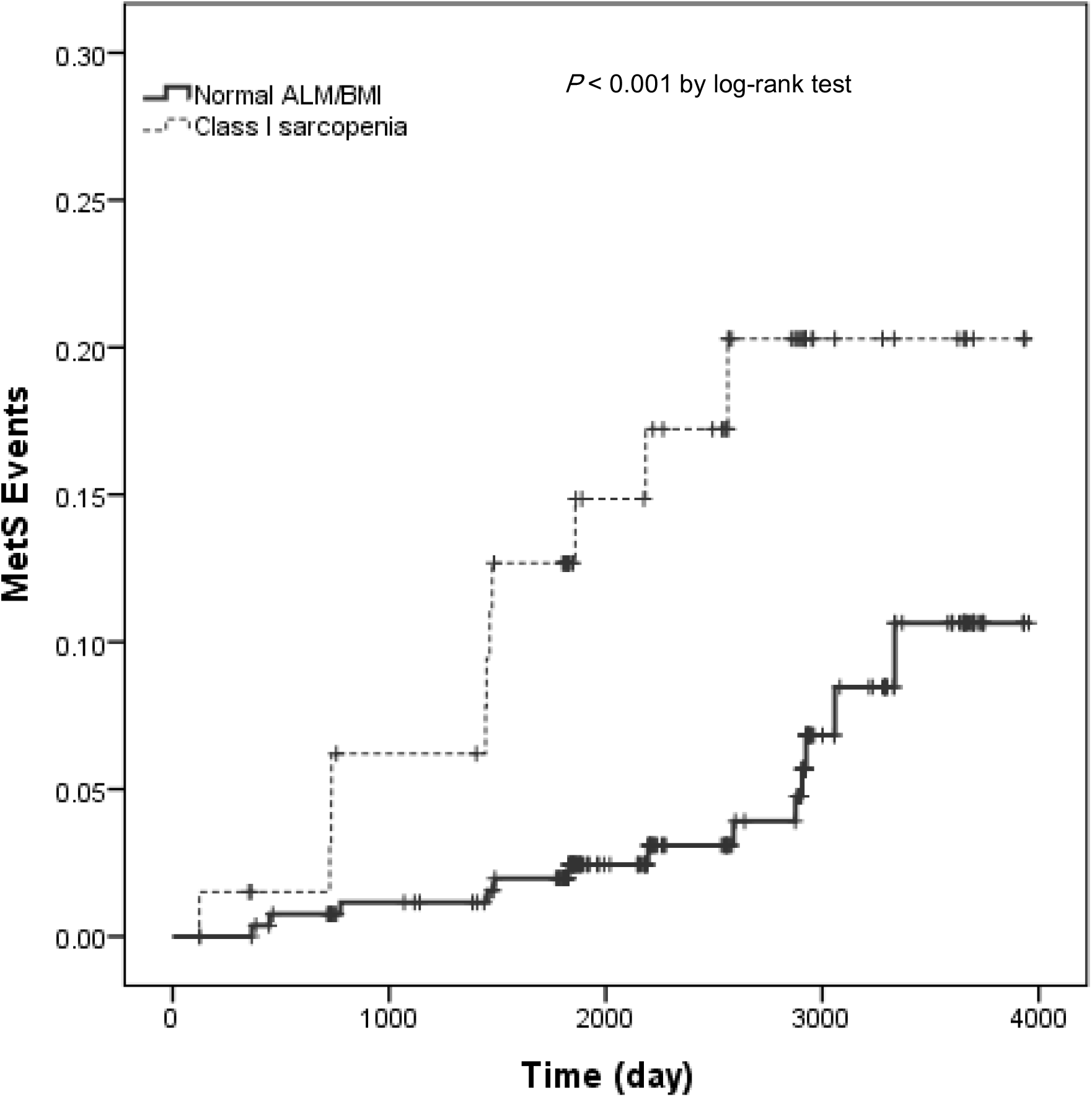
Kaplan-Meier curves for events of incident metabolic syndrome (MetS) according to baseline ALM/BMI. Bold line shows normal ALM/BMI, and dashed line shows Class I sarcopenia.

**Fig. 3.**
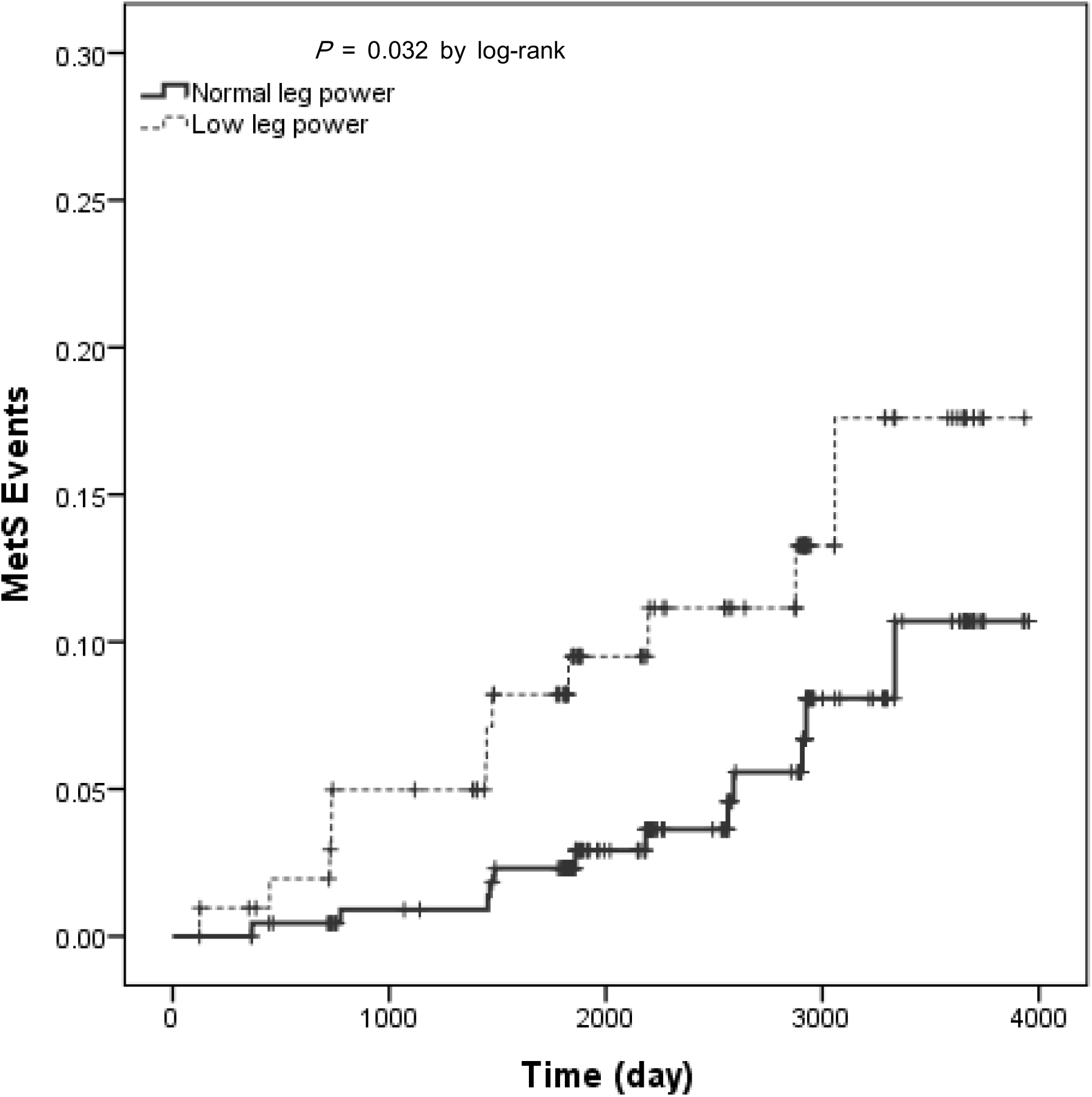
Kaplan-Meier curves for events of incident metabolic syndrome (MetS) according to baseline leg power (W/kg). Bold line shows normal leg power, and dashed line shows low leg power.

Cox proportional hazard regression analyses were performed in Table 2. We found that the subjects with Class I sarcopenia defined by SMI or ALM/BMI was significantly associated with an increased adjusted hazard ratio (AHR) for incident MetS (3.22, 95%CI 1.25-8.27 for SMI, and 3.19, 95%CI 1.31-7.74 for ALM/BMI) compared with the subjects with normal relative skeletal muscle mass, after adjusting age, family history of diabetes, smoking status, and physical activity. In contrast, for the leg power per body weight, the incidence of MetS was not significantly different between low leg power and normal leg power groups after adjusting those variables (AHR = 1.23, 95%CI; 0.50-3.04).

**Table 2.** Association between baseline relative muscle mass or power and incidence of metabolic syndrome (Cox model) (N = 346)

|  | Model 1 | Model 2 | Model 3 |
| --- | --- | --- | --- |
|  | HR (95%CI) | HR (95%CI) | HR (95%CI) |
| Normal SMI | 1 (ref) | 1 (ref) | 1 (ref) |
| Class I Sarcopenia | 2.61 (1.14-5.98)<br>P = 0.023 | 3.16 (1.35-7.39)<br>P = 0.008 | 3.22 (1.25-8.27)<br>P = 0.015 |
| Normal ALM/BMI | 1 (ref) | 1 (ref) | 1 (ref) |
| Class I Sarcopenia | 3.88 (1.74-8.68)<br>P = 0.001 | 3.52 (1.57-7.92)<br>P = 0.002 | 3.19 (1.31-7.74)<br>P = 0.010 |
| Normal leg power | 1 (ref) | 1 (ref) | 1 (ref) |
| Low leg power | 2.34 (1.05-5.22)<br>P = 0.037 | 1.67 (0.74-3.84)<br>P = 0.214 | 1.23 (0.50-3.04)<br>P = 0.654 |

SMI, skeletal muscle mass index; BMI, body mass index; ALM, appendicular lean mass; HR, hazard ratio; CI, confidence interval.
Model 1: crude
Model 2: Model 1 + further adjusted for age
Model 3: Model 2 + further adjusted for family history of diabetes, smoking status, physical activity

## 4. Discussion

The major findings of this 7-year prospective study was subjects with Class I sarcopenia defined by SMI (ALM/weight × 100%) or ALM/BMI was significantly associated with an increased adjusted HR for incident MetS compared with the subjects with normal skeletal muscle mass, after adjusting age, family history of diabetes, smoking status, and physical activity. In contrast, the incidence of MetS was not significantly different between low leg power and normal leg power groups after adjusting those variables.

Skeletal muscle mass (SMM) is strongly correlated with body size [20]. Thus, EWGSOP2 stated that “when quantifying muscle mass, the absolute level of SMM or ALM can be adjusted for body size in different ways, namely using height squared (ALM/height^2^), weight (ALM/weight) or body mass index (ALM/BMI) [2].” Preferred adjustment has been a subject of debate. Janssen et al. indicated that SMM/weight is associated with functional impairment and disability in NHANES III participants aged 18 and older [21]. Janssen also indicated that SMM/height^2^ is associated with physical disability in the subjects aged 65 and older in Cardiovascular Health Study (CHS) database [22]. Furushima et al. indicated low ALM/height^2^ is associated with bone mineral density but not with MetS variables, and ALM/weight is associated with MetS variables but not with bone mineral density [13]. Many previous studies also indicated that ALM/weight is associated with MetS [9-13]. A recent study found the significant association between low ALM/weight and MetS development in a 7-year retrospective study [23]. The result of the current study is consistent with those cross-sectional studies or the retrospective study, and this is the first prospective study to examine the association of low ALM/weight or ALM/BMI and MetS development, to the best of our knowledge. On the contrary, high ALM/height^2^ or high absolute ALM had significantly higher incident of MetS during the follow-up period. This is consistent with the recent cross-sectional study [24] and previous studies [7-14]. It is because the people who have higher body weight tend have higher muscle mass. Therefore, the normalization of muscle mass by body weight or BMI is needed to examine the association between SMM and MetS.

The association between muscle strength or power and MetS has also been examined. Jurca et al. indicated that low muscular strength index computed by combining the one-repetition maximum score for the bench press and the leg press expressed as weight lifted per kilogram body weight was significantly associated with high prevalence of MetS [7, 8]. Very recently, Zhang et al. examined the association between the prevalence of MetS and absolute or relative values of muscle strength in women [14]. They concluded that the prevalence of MetS increased with low relative grip and leg strengths (per kilogram body weight). Conversely, low absolute muscle strength was associated with low MetS prevalence [14]. The current longitudinal study showed the significant association between low relative muscle power and the development of MetS, but the association was no more significant after adjusting age, family history of diabetes, smoking status, and physical activity.

Several mechanisms may affect the association between low relative skeletal muscle mass and development of MetS, including physical inactivity, malnutrition, insulin resistance, inflammation, and myokines [11]. Skeletal muscle is the main site of glucose uptake and utilization. Low relative skeletal muscle mass may increase insulin resistance and thereby induce MetS. Actually, the HOMA-IR was negatively associated with low relative muscle mass.

This study has several limitations. First, because of limited sample size, we could not test many adjusting variables in this study. There may be possible cofounders between low relative skeletal muscle mass and development of MetS. Second, the muscle quality and composition, such as fat infiltration [25, 26], fibrosis [27, 28], relative expansion of extracellular compartments [3, 29, 30], are important issue to assess muscle tissues. However, we could only assess ASM in this study. Further studies are needed to address this issue. Third, recently, BIA is used many studies to assess ASM, but the BIA method is a secondary indirect method to estimate body composition [31]. BIA is possibly influenced by edema, exercise, circadian and seasonal variations [32-34]. Although BIA was measured in the morning without any exercise and fasting state in this study, still seasonal variations may affect the current results.

## 5. Conclusions

In conclusion, our results show the relative skeletal muscle mass against body weight (ALM/weight) or body mass index (ALM/BMI) is negatively associated the future development of MetS. The relative skeletal muscle mass was negatively associated with HOMA-IR. To maintain the relative skeletal muscle mass is important to prevent developing MetS in Japanese women.

## Data Availability

Data cannot be shared publicly because of Guideline of the Ethics Committee

## Funding

The study was funded by the Ministry of Health, Labour and Welfare (Health and Labour Sciences Research Grant: 200825016B and 201222028B).

## Acknowledgments

The authors thank Dr. Kumpei Tanisawa, Dr. Harumi Ohno, Dr. Kana Konishi, Dr. Michiya Tanimoto, Dr. Noriko Tanaka, Dr. Hiroshi Kawano, Dr. Kenta Yamamoto, Dr. Motoyuki Iemitsu, Ms. Azusa Sasaki, Ms. Yumi Ohmori, Ms. Rie Katayama, Mr. Zhenbo Cao, Ms. Eriko Kubo, Ms. Miyuki Hayashi, Mr. Satoshi Hanawa, Ms. Naeko Kurose, Ms. Aiko Hirosako, Ms. Sayaka Nakamura, Ms. Hidemi Hara, Ms. Miki Yoshida, Mr. Satoshi Kurita, Ms. Noriko Wada, Ms. Miho Okamoto, Ms. Hisako Ito, Ms. Kinue Nakajima, Ms. Kaori Sato, Ms. Akie Morishita, and Ms. Kazumi Kajiwara, who significantly contributed to the realization of this study through their long-term involvement as researchers or research assistants. Moreover, the authors would like to express their gratitude to all the subjects who participated in the study and to all research professionals involved in the NEXIS protocol.

## Conflicts of Interest

There is no conflicts of interest in this study.

## Notes

### Competing Interest Statement

The authors have declared no competing interest.

### Funding Statement

The funders had no role in study design, data collection and analysis, decision to publish, or preparation of the manuscript.

### Author Declarations

All procedures were reviewed and approved by the ethics committees of the National Institutes of Biomedical Innovation, Health and Nutrition (6008, Kenei14-02). All participants provided written consent for participation in the study.

